# Preterm birth rates during the COVID-19 lockdown in Queensland Australia

**DOI:** 10.1101/2021.08.06.21261711

**Authors:** Brittany Jasper, Tereza Stillerova, Christopher Anstey, Edward Weaver

## Abstract

**Background:** Preventative strategies for preterm birth are lacking. Recent evidence proposed COVID-19 lockdowns may have contributed to changes in preterm birth. We aimed to determine the prevalence of preterm birth during the COVID-19 lockdown on the Sunshine Coast and in the state of Queensland, Australia.

**Methods:** Retrospective cohort analysis of all births in Queensland including the Sunshine Coast University Hospital, during two epochs, April 1–May 31, 2020 (lockdown) and June 1–July 31, 2020 (post-lockdown), compared to antecedent calendar-matched periods in 2018–2019. Prevalence of preterm birth, stillbirth, and late terminations were examined.

**Results:** There were 64,989 births in Queensland from April to July 2018–2020. At the Sunshine Coast University Hospital, there was a significantly higher chance of birth at term during both lockdown (OR 1.81, 95% CI 1.17, 2.79; P=0.007) and post-lockdown (OR 2.01, 95% CI 1.27, 3.18; P=0.003). At the same centre, prevalence of preterm birth was 5.5% (30/547) during lockdown, compared to 9.1% (100/1095) in previous years, a 40.0% relative reduction (P=0.016). At this centre during lockdown, emergency caesareans concurrently decreased (P=0.034) and vacuum-assisted births increased (P=0.036). In Queensland overall, there was a nonsignificant decrease in the prevalence of preterm birth during lockdown.

**Conclusions:** There is a link between lockdown and a reduction in the prevalence of preterm birth on the Sunshine Coast. The cause is speculative. Further research is needed to determine a causal link and assess if this can be translated into management which provides a sustained reduction in preterm birth and its associated morbidity and mortality.

## INTRODUCTION

Preterm birth (PTB) is a major problem worldwide and is the leading cause of death in children under five.[1] Despite advances in perinatal care, there has been no sustained reduction in the prevalence of PTB and preventative measures remain only partially successful.[2]

The World Health Organisation (WHO) declared COVID-19 a pandemic on 11 March, 2020.[3] This was followed by lockdowns across the globe, causing unprecedented effects on society and human behaviour in ways not seen before. In Queensland, Australia, the restrictions to mitigate the spread of COVID-19 culminated in a nine-week state-wide lockdown from March 30 to June 1, 2020, with a phased easing of restrictions.[4] During lockdown, all shops, restaurants, schools, universities, and non-essential workplaces were closed.[4-5] Residents were prohibited from leaving their homes except for essential reasons (*Figure 1*).[4]

**Figure 1.**
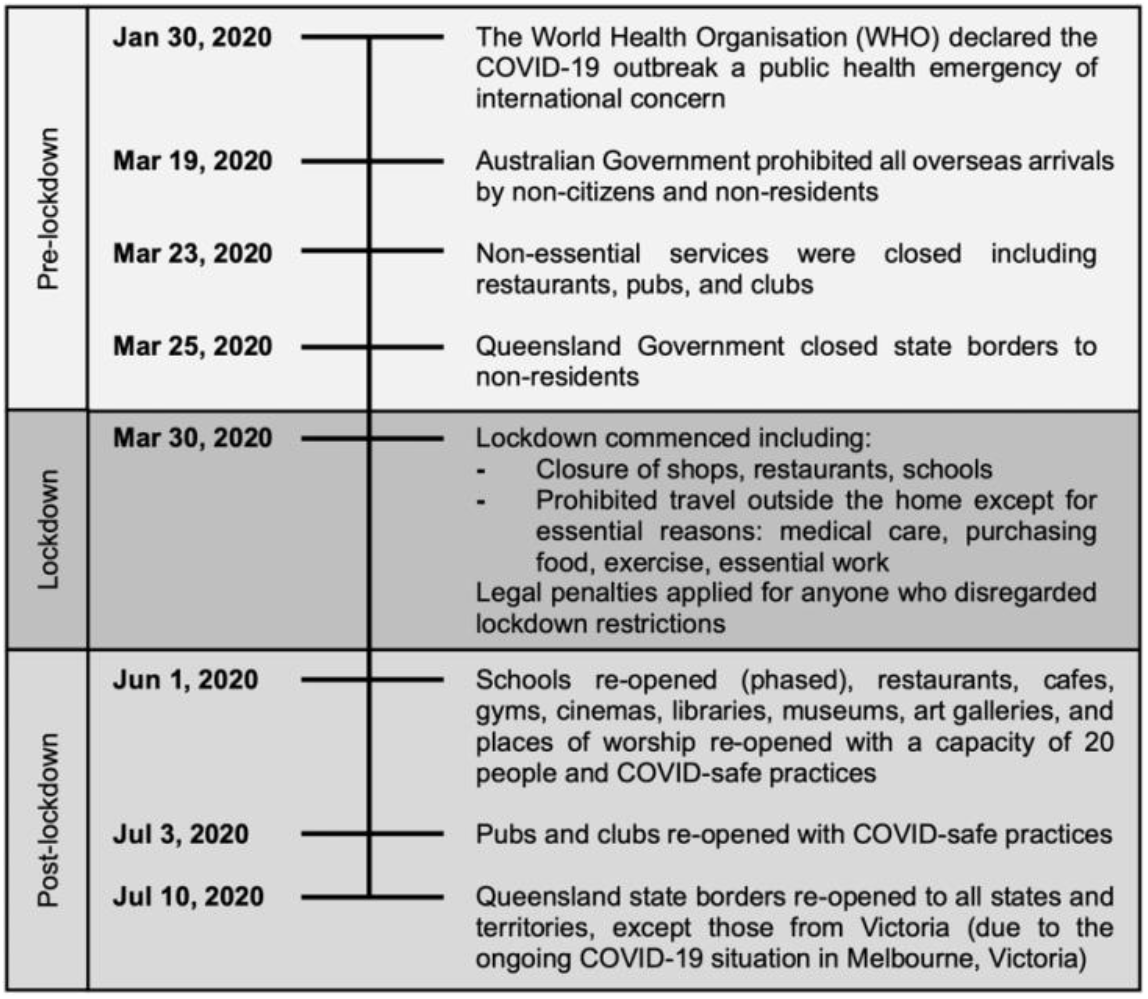
Timeline of COVID-19 restriction implementation in Queensland, Australia.[4,5]

To minimise the risk of virus transmission, models of antenatal care at the Sunshine Coast University Hospital (SCUH) were adapted.[6] Women were risk-triaged by senior obstetric staff to either virtual or in-person appointments utilising COVID-19 safe practices.[6]

In the wake of the pandemic, research emerged demonstrating the varied impact of lockdown on the prevalence of PTB worldwide.[7-16] We aimed to determine the prevalence of PTB during the COVID-19 lockdown on the Sunshine Coast and in the state of Queensland, Australia.

## METHODS

### Study population

This study focused on pregnant women in two geographical locations in Australia. Firstly, the state of Queensland, which services the health needs of over 5 million people, and secondly, SCUH, a health service which provides care for the 400,000 people living in this area of Queensland.[17-18] Of pregnant women in Queensland, 6.8% reside in the SCUH catchment.[19] Three percent of pregnant women in Queensland were aged less than 20 years and four percent were aged over 40 years[19], 7.5% identified as Aboriginal or Torres Strait Islander[19], 21.0% were obese at the time of conception[20], and 11% smoked during pregnancy[20].

### Ethics approval and data collection

This study was reviewed by the Prince Charles Hospital Human Research Ethics Committee and approval was granted as a low or negligible risk project (Project ID 70040; November 3, 2020). Data pertaining to SCUH was collected from the health service birth registry. Queensland state-wide data was sourced from The Perinatal Data Collection, Department of Health.

### Study design

Retrospective cohort analysis of all births in Queensland, Australia was carried out during two epochs, April 1–May 31, 2020 (lockdown) and June 1–July 31, 2020 (post-lockdown), compared to antecedent calendar-matched periods from the preceding two years 2018–2019. The prevalence of PTB, stillbirth, and late terminations were examined. Inclusion criteria were all births after 20 weeks’ gestation (live and still) and late terminations. All PTB defined as less than 37 weeks’ gestation were analysed, including sub-group analyses of extreme preterm (20+0–27+6 weeks’ gestation), very preterm (28+0– 31+6 weeks’ gestation), and moderate to late preterm (32+0–36+6 weeks’ gestation). To facilitate comparison of birth data at SCUH to the state of Queensland overall, and to wholly represent all births after 20 weeks’ gestation, no births were excluded because of multiple births, stillbirths after 20 weeks’ gestation, or terminations after 20 weeks’ gestation. At SCUH, perinatal characteristics were collected, including maternal age, body mass index (BMI), parity, mode of delivery, and infant sex. Further analysis of PTB data at SCUH was carried out to examine the proposed aetiology of PTB and birth outcomes including spontaneous live PTB, iatrogenic live PTB, stillbirth after 20 weeks’ gestation, and late termination after 20 weeks’ gestation. We subclassified spontaneous live PTB aetiologies into preterm premature rupture of membranes, preterm labour and birth, and multiple pregnancy. Subclassifications for iatrogenic (medically indicated) live PTB included intrauterine growth restriction, hypertension (including pre-eclampsia and HELLP syndrome (haemolysis, elevated liver enzymes, low platelet count)), multiple pregnancy, fetal distress, placental abruption and antepartum haemorrhage, and other maternal comorbidities.

### Statistical analysis

All data was anonymised and deidentified prior to analysis. Univariate logistic regression was performed to identify potential predictors of preterm or term birth. All variables with *P*<0.20 were included in multivariate logistic regression analysis. The prevalence of PTB, stillbirth, and late terminations were calculated. Percent change was calculated to assess for any difference in prevalence between time periods. Descriptive statistics were reported as mean and standard deviation (SD) for normally distributed continuous data, median and interquartile range (IQR) for non-normally distributed data, and frequencies and percentages for categorical data. Continuous data were tested for normality using the Shapiro-Wilk test and analysed using the t-test. Categorical data were analysed using either a Mann-Whitney U-test, Chi-squared, or Fisher’s exact test, where appropriate. P-values <0.05 were considered significant. All analyses were conducted using R statistical software.

### Patient and public involvement

Patient and public involvement began in the early stages of project planning. The Preterm Infants Parents Association (PIPA) is a registered Australian charity and support network for families with premature babies.[21] The PIPA team reviewed the proposed study design, research questions, and confirmed all data collection processes were confidential. Once the study is published, a plain language summary will be distributed to the PIPA database of preterm infants’ parents and their network.

## RESULTS

There were 64,989 births in the state of Queensland, Australia, including 3,296 births at SCUH between April 1–July 31, 2018–2020. Univariate logistic regression of births at SCUH revealed maternal age, BMI, parity, and infant sex were all nonsignificant predictors of PTB. Multivariate logistic regression of births at SCUH demonstrated a significantly higher chance of birth at term during both the lockdown period April–May 2020 (OR 1.81, 95% CI 1.17, 2.79; P=0.007) and the post-lockdown period June–July 2020 (OR 2.01, 95% CI 1.27, 3.18; P=0.003), when compared with antecedent calendar-matched periods in 2018–2019 (*supplementary material*).

All births at SCUH and in the state of Queensland overall are summarised in *table 1*. During lockdown, the prevalence of PTB at SCUH was 5.5% (30/547), compared to 9.1% (100/1095) during the same period in previous years, representing a 40.0% relative reduction in PTB (P=0.016). This reduction in PTB at SCUH appeared to be driven by moderate to late PTB (32+0–36+6 weeks’). The prevalence of moderate to late PTB was 4.6% (25/547) during lockdown compared to 7.5% (82/1095) in previous years, a 39.0% relative reduction (P=0.034). In contrast, the prevalence of PTB in the state of Queensland overall was 8.6% (870/10154) during lockdown compared to 8.9% (1871/20939) in previous years, a nonsignificant 4.1% relative reduction in PTB (P=0.33).

**Table 1.** Birth outcomes during the lockdown and post-lockdown periods from 2018–2020 at the Sunshine Coast University Hospital and in the state of Queensland overall.

| Birth outcome<br>No./Total No. (%) | No lockdown<br>Apr–May 2018–9<br>(n=22 034) | Lockdown<br>Apr–May 2020<br>(n=10 701) | P | No lockdown<br>Jun–Jul 2018–9<br>(n=21 568) | Post-lockdown<br>Jun–Jul 2020<br>(n=10 686) | P |
| --- | --- | --- | --- | --- | --- | --- |
| **Total preterm births/Total births** |  |  |  |  |  |  |
| SCUH† | 100/1095 (9.13) | 30/547 (5.48) | **0.016** | 80/1130 (7.08) | 26/524 (4.96) | 0.12 |
| QLD‡ | 1871/20939 (8.94) | 870/10154 (8.57) | 0.33 | 1865/20438 (9.13) | 921/10162 (9.06) | 0.87 |
| **Extreme preterm (20+0–27+6)** |  |  |  |  |  |  |
| SCUH | 12/1095 (1.10) | 3/547 (0.55) | 0.41 | 5/1130 (0.44) | 5/524 (0.95) | 0.21 |
| QLD | 193/20939 (0.92) | 113/10154 (1.11) | 0.11 | 166/20438 (0.81) | 87/10162 (0.86) | 0.69 |
| **Very preterm (28+0–31+6)** |  |  |  |  |  |  |
| SCUH | 6/1095 (0.55) | 2/547 (0.37) | >0.99 | 3/1130 (0.27) | 1/524 (0.19) | >0.99 |
| QLD | 163/20939 (0.78) | 79/10154 (0.78) | >0.99 | 191/20438 (0.93) | 77/10162 (0.76) | 0.12 |
| **Moderate/late preterm (32+0–36+6)** |  |  |  |  |  |  |
| SCUH | 82/1095 (7.49) | 25/547 (4.57) | **0.034** | 72/1130 (6.37) | 20/524 (3.82) | **0.045** |
| QLD | 1515/20939 (7.24) | 678/10154 (6.68) | 0.09 | 1508/20438 (7.38) | 757/10162 (7.45) | 0.84 |

†SCUH: Sunshine Coast University Hospital
‡QLD: State of Queensland, Australia
**Bold** indicates significance of P value <0.05

After lockdown restrictions eased, the prevalence of PTB at SCUH was 5.0% (26/524) during the post-lockdown period compared to 7.1% (80/1130) in previous years, a nonsignificant 29.9% relative reduction in PTB (P=0.12). This decrease appeared to be driven by moderate to late PTB, as the prevalence in this group was 3.8% (20/524) during post-lockdown, compared to 6.4% (72/1130) in previous years, a 40.0% relative reduction in moderate to late PTB post-lockdown (P=0.045). In the state of Queensland overall, there was no significant difference in PTB observed in the post-lockdown period compared to previous years.

*Table 2* outlines the perinatal characteristics of births at SCUH between April 1–July 31, 2018 to 2020. Most pregnant women were aged 20–40 years and only 4.3% of women were in the higher risk age groups for PTB of ≤20 years or ≥40 years.[22] There was a general increase in BMI in both the lockdown and post-lockdown groups in 2020 when compared to 2018–2019, consistent with the rising prevalence of obesity.[6] At SCUH during lockdown, we observed a 29.4% relative decrease in emergency caesarean sections (in labour) and concurrent 41.7% relative increase in vacuum-assisted births when compared to previous years (P=0.034, P=0.036, respectively).

**Table 2.** Perinatal characteristics at the Sunshine Coast University Hospital from 2018–2020.

| Characteristic<br>No./Total No. (%) | No lockdown<br>Apr–May 2018–9 | Lockdown<br>Apr–May 2020 | P | No lockdown<br>Jun–Jul 2018–9 | Post-lockdown<br>Jun–Jul 2020 | P |
| --- | --- | --- | --- | --- | --- | --- |
| **Maternal age<sup>§</sup> (median, IQR)** | 31 (27–34) | 30 (26.5–34) | >0.99 | 30 (26–34) | 31 (27–34) | 0.78 |
| Age under 20 years | 23/1095 (2.10) | 8/547 (1.46) | 0.40 | 15/1130 (1.33) | 4/524 (0.76) | 0.46 |
| Age 20 to 40 years | 1045/1095 (95.43) | 527/547 (96.35) | 0.90 | 1078/1130 (95.40) | 505/524 (96.38) | 0.89 |
| Age over 40 years | 27/1095 (2.47) | 12/547 (2.19) | 0.74 | 37/1130 (3.27) | 15/524 (2.86) | 0.67 |
| **Body Mass Index<sup>§</sup> (median, IQR)** | 27 (24–58) | 29 (25–54) | **<0.01** | 28 (24–60) | 29 (25–58) | **0.030** |
| Underweight (<18) | 30/1095 (2.74) | 10/547 (1.83) | 0.27 | 25/1130 (2.21) | 3/524 (0.57) | **0.022** |
| Healthy weight (18–25) | 603/1095 (55.07) | 234/547 (42.78) | **<0.01** | 570/1130 (50.45) | 242/524 (46.18) | 0.34 |
| Overweight (25–30) | 296/1095 (27.03) | 165/547 (30.16) | 0.32 | 304/1130 (26.90) | 150/524 (28.63) | 0.58 |
| Obese (>30) | 142/1095 (12.97) | 113/547 (20.66) | **<0.01** | 194/1130 (17.17) | 98/524 (18.70) | 0.52 |
| Unknown | 24/1095 (2.19) | 25/547 (4.57) | **<0.01** | 37/1130 (3.27) | 31/524 (5.92) | **0.016** |
| **Parity** |  |  |  |  |  |  |
| Nulliparous | 425/1095 (38.81) | 218/547 (39.85) | 0.79 | 448/1130 (39.64) | 202/524 (38.55) | 0.78 |
| Primiparous | 384/1095 (35.07) | 190/547 (34.74) | 0.93 | 375/1130 (33.19) | 183/524 (34.92) | 0.63 |
| Multiparous | 286/1095 (26.12) | 139/547 (25.41) | 0.81 | 307/1130 (27.17) | 139/524 (26.53) | 0.84 |
| **Mode of delivery** |  |  |  |  |  |  |
| Spontaneous vaginal birth | 661/1095 (60.37) | 318/547 (58.14) | 0.66 | 651/1130 (57.61) | 278/524 (53.05) | 0.35 |
| Vacuum-assisted birth | 96/1095 (8.77) | 68/547 (12.43) | **0.036** | 143/1130 (12.65) | 69/524 (13.17) | 0.80 |
| Forceps-assisted birth | 19/1095 (1.74) | 14/547 (2.56) | 0.27 | 23/1130 (2.04) | 16/524 (3.05) | 0.22 |
| Vacuum + forceps-assisted birth | 1/1095 (0.09) | 0/547 (0.00) | >0.99 | 1/1130 (0.09) | 0/524 (0.00) | >0.99 |
| Elective caesarean | 125/1095 (11.41) | 79/547 (14.44) | 0.12 | 136/1130 (12.04) | 72/524 (13.74) | 0.39 |
| Emergency caesarean (in labour) | 156/1095 (14.24) | 55/547 (10.05) | **0.034** | 152/1130 (13.45) | 79/524 (15.08) | 0.44 |
| Emergency caesarean (no labour) | 37/1095 (3.38) | 13/547 (2.38) | 0.28 | 24/1130 (2.12) | 10/524 (1.91) | 0.78 |

<sup>§</sup>P values calculated with the Mann-Whitney U test. Unmarked P values calculated using Chi-squared or Fisher's exact test, where appropriate.
**Bold** indicates significance of P value <0.05

Upon further analysis of PTB data at SCUH, no significant change was identified in either spontaneous or iatrogenic (medically indicated) live PTB during lockdown or post-lockdown (*table 3*). A nonsignificant increase in stillbirth was observed during lockdown and post-lockdown when compared to previous years at SCUH, although the numbers were very low (P=0.09, P=0.08, respectively). There was no change in the prevalence of late terminations, compared to previous years at SCUH.

**Table 3.** Birth outcomes and proposed aetiologies for preterm birth during the lockdown and post-lockdown periods from 2018–2020 at the Sunshine Coast University Hospital.

| Birth outcome<br>No./Total No. (%) | No lockdown<br>Apr–May 2018–9 | Lockdown<br>Apr–May 2020 | P | No lockdown<br>Jun–Jul 2018–9 | Post-lockdown<br>Jun–Jul 2020 | P |
| --- | --- | --- | --- | --- | --- | --- |
| **Spontaneous live preterm birth** | 52/100 (52.00) | 16/30 (53.33) | 0.94 | 50/80 (62.50) | 11/26 (42.31) | 0.33 |
| Preterm premature rupture of membranes | 26/100 (50.00) | 8/30 (26.67) | 0.96 | 28/80 (35.00) | 7/26 (26.92) | 0.58 |
| Preterm labour and birth | 16/100 (30.77) | 6/30 (20.00) | 0.67 | 18/80 (22.50) | 4/26 (15.38) | 0.78 |
| Multiple pregnancy | 10/100 (19.23) | 2/30 (6.67) | >0.99 | 4/80 (5.00) | 0/26 (0.00) | 0.57 |
| **Iatrogenic live preterm birth** | 39/100 (39.00) | 11/30 (36.67) | 0.88 | 24/80 (30.00) | 8/26 (30.77) | 0.96 |
| Intrauterine growth restriction | 10/100 (10.00) | 3/30 (10.00) | >0.99 | 4/80 (5.00) | 1/26 (3.85) | >0.99 |
| Hypertension | 8/100 (8.00) | 2/30 (6.67) | >0.99 | 5/80 (6.25) | 2/26 (7.69) | >0.99 |
| Multiple pregnancy | 17/100 (17.00) | 4/30 (13.33) | 0.79 | 10/80 (12.50) | 4/26 (15.38) | 0.75 |
| Other maternal comorbidities | 2/100 (2.00) | 1/30 (3.33) | 0.55 | 2/80 (2.50) | 1/26 (3.85) | >0.99 |
| Fetal distress | 1/100 (1.00) | 1/30 (3.33) | 0.42 | 2/80 (2.50) | 0/26 (0.00) | >0.99 |
| Placental abruption or antepartum haemorrhage | 1/100 (1.00) | 0/30 (0.00) | >0.99 | 1/80 (1.25) | 0/26 (0.00) | >0.99 |
| Stillbirth ≥20 weeks | 2/100 (2.00) | 3/30 (10.00) | 0.09 | 3/80 (3.75) | 4/26 (15.38) | 0.08 |
| Termination ≥20 weeks | 7/100 (7.00) | 0/30 (0.00) | 0.35 | 3/80 (3.75) | 3/26 (11.54) | 0.18 |

**Bold** indicates significance of P value <0.05

## DISCUSSION

Australia enforced strict lockdowns to mitigate the spread of COVID-19, reaching a stringency level of 71.3 in a 1 to 100 scale on the Oxford Government Response Tracker.[23] Residents were not permitted to leave their home except for essential reasons.[4] At the Sunshine Coast University Hospital (SCUH) in Queensland Australia, there was a significantly higher chance of birth at term during both the lockdown and post-lockdown periods, when compared to previous years. During lockdown, a 40.0% relative reduction in the prevalence of PTB was observed at SCUH (*table 1*). This finding is consistent with similar studies worldwide which reported a significantly lower prevalence of PTB during lockdown.[7,9,10,13,14] In contrast to the reduction in PTB observed at SCUH, there was a nonsignificant decrease in PTB observed in the state of Queensland overall during lockdown. On the Sunshine Coast and in Brisbane[9], both cities in Queensland, there was a reduction in PTB, however in our study, this reduction was not observed for the state overall. It is possible that the effect of lockdown on PTB was more apparent in cities, although this requires further investigation. At SCUH, the reduction in the prevalence of PTB during lockdown was driven by moderate to late PTB, in line with similar evidence from Australia.[9] This reduction in moderate to late PTB was sustained during the post-lockdown period, suggesting the possibility that altered maternal behaviours enforced during lockdown may have persisted even after restrictions were lifted.

The obstetric antecedents of PTB include delivery for materno-fetal indications (iatrogenic), spontaneous preterm labour and birth, or preterm premature rupture of membranes.[22] In high-income countries such as Australia, 30% of PTB are iatrogenic, while 70% are spontaneous.[2] The aetiology of most spontaneous PTB is unknown, although in singleton pregnancies the most common cause is likely intrauterine inflammation and placental anomalies.[2] We did not observe a significant difference in the prevalence of spontaneous or iatrogenic live PTB during lockdown or post-lockdown, compared to previous years. However, other studies from Australia[9] and Israel[15] reported a reduction in iatrogenic PTB during lockdown.

Although differences were observed in the prevalence of PTB, the cause for this is speculative. During lockdown, maternal behaviours were changed as a result of lockdown and public health messaging, and women likely had increased health awareness. During lockdown, Australia noted improved air quality[24], which is known to impact PTB.[25] Although the causality between PTB and work remains controversial, with more women working from home or not working, there may have been more time for sleep, exercise, and healthy eating.[16] However, it is important to recognise the known negative impacts of lockdown for some women, as there was a rise in domestic violence and poor mental health in Australia during lockdown.[26-27]

When postulating factors which may have contributed to the reduction in PTB, it is reasonable to consider a decrease in the transmission of infections, such as influenza, a known risk factor for PTB.[28-29] Pregnant women have an increased risk of severe influenza, and severe infection increases the risk of PTB and stillbirth.[28-29] It is recommended that pregnant women receive the influenza vaccine antenatally, which reduces the risk of influenza for the pregnant woman, the risk of PTB, and confers immunity to the newborn for three to six months.[29-30]

In Queensland, Australia, maternal influenza vaccination uptake has been suboptimal with 42.0% of women receiving the vaccine in 2018, despite it being encouraged and free of charge.[20] In the context of COVID-19, the Australian government introduced measures to increase the uptake of influenza vaccination and emphasised the importance of pregnant women receiving the vaccine.[31-32] The time to immunity after influenza vaccination is approximately 2 weeks.[33] Influenza vaccination uptake in Australia increased from 3.5 million in 2018 to 7.3 million doses by May 2020 during lockdown, which may be attributed to effective public health messaging and increased health awareness during the pandemic.[34] In 2020, Australia noted the lowest rates of influenza in the past decade, almost 8 times lower than the five-year average[35], likely as a result of exceptionally high vaccination uptake and reduced transmission of infections during lockdown. Both high influenza vaccination uptake and low influenza infection rates may have contributed to the reduction observed in PTB, although this requires further study.

During the lockdown period at SCUH, there was a 29.4% relative decrease in emergency caesarean sections and concurrent 41.7% relative increase in vacuum-assisted vaginal births *(table 2)*. This trend did not extend into the post-lockdown period. It is possible that reduced caesarean sections during lockdown were due to protracted COVID-related infection control policies, potential altered staff levels with some staff affected by or shielding from COVID, or other factors not yet delineated. A similar study from Japan demonstrated a comparable reduction in caesarean sections during the lockdown.[13] However, a British study[11] showed no change, and an Indian study[12] showed an increase in caesarean sections during lockdown. Further research into provision of maternity care during the lockdown was beyond the scope of this study.

At SCUH, there was a nonsignificant increase in stillbirth during lockdown and post-lockdown *(table 3)*. Other studies demonstrated a wide variation in stillbirth rates during lockdown; studies from Australia[9,14], England[36-37], and Israel[15] showed no change in stillbirth, while other studies from England[11], India[12], and Italy[8] demonstrated an increase in stillbirth rates during lockdown. This conflicting evidence underscores the need for ongoing research into stillbirth during the COVID-19 pandemic.

To our knowledge, our study is the first to examine state-wide PTB data during the COVID-19 pandemic in Australia. This allowed for comparison of one health region to a large state overall, including metropolitan, regional, and rural areas. This paper is one of only three studies from Australia to investigate the prevalence of PTB during the pandemic. For births at SCUH, we analysed perinatal characteristics including maternal demographics, mode of delivery, maternal comorbidities, and birth outcomes such as spontaneous live PTB, iatrogenic live PTB, stillbirth, and late terminations.

Our study has limitations, which should be considered when interpreting results. As this study was observational, it was not possible to account for unmeasured confounding variables. Although detailed data was collected on all births at SCUH, state-wide data for Queensland was limited to birth outcomes and gestational age, as per the predetermined perinatal data collection format. As our health service opened a new tertiary hospital in April 2017 (SCUH), retrospective analysis was only completed for two years prior from 2018–2019. We acknowledge that longer retrospective analysis would be preferable to further compare year-by-year variations in birth outcomes.

Further research is required from ethnically, geographically, and socioeconomically diverse regions and high-risk groups for PTB such as Aboriginal and Torres Strait Islander women in Australia. Qualitative analysis of maternal behaviours during lockdown was beyond the scope of this study but would be highly informative.

## Conclusion

There is a link between lockdown and a reduction in the prevalence of PTB on the Sunshine Coast. The cause is speculative at present. Further research is needed to determine a causal link and assess if this can be translated into management which provides a sustained reduction in PTB and its associated morbidity and mortality.

## Data Availability

The authors confirm that the data supporting the findings of this study are available within the article and its supplementary materials.

## Acknowledgements

The authors would like to acknowledge the Gubbi Gubbi people, the traditional owners of the land on the Sunshine Coast, and pay respect to Elders past, present, and emerging. We thank our obstetric, midwifery, neonatal, and administrative colleagues at SCUH who supported this project, particularly Carol Newell, Nikki Doogue, Tracy Kerr, and Tammy Doyle for collation of obstetric data. The Perinatal Data Collection, Queensland Department of Health are acknowledged state-wide data collection. We acknowledge the Queensland Facility for Advanced Bioinformatics, Nicholas Matigan for assistance with statistical analyses and Nicholas Snels for his comments. PIPA and Eileen Cooke are gratefully acknowledged for their independent review of our study design.

## Contributors

BJ, TS, EW conceptualised and designed the study. BJ and TS obtained and analysed the initial data. BJ, TS, CA, EW analysed the final data. BJ and TS drafted the initial manuscript. BJ, TS, CA, EW critically reviewed and revised the final manuscript. EW supervised the research project. All authors approved the final manuscript as submitted. All authors listed meet authorship criteria.

## Funding

The authors did not receive any funding or financial sponsorship to carry out this research.

## Competing interests

None declared.

## Ethics approval

As per the National Health and Medical Research Council of Australia (NHMRC), this study was reviewed by The Prince Charles Hospital Human Research Ethics Committee and approval was granted as a low or negligible risk project (Project ID 70040; November 3, 2020).

## Disclaimer

The views and postulations expressed in this paper are those of the authors, and are not representative of any employing agencies such as the Sunshine Coast University Hospital, Griffith University, the University of Queensland, North West Anglia Healthcare NHS Trust, The Perinatal Data Collection, Queensland Government, Department of Health, or the Preterm Infants Parents Association.

## Patient and public involvement

A public group (PIPA) were consulted in this study, see methods.

## What is already known on this topic

- In light of the COVID-19 pandemic, international research has demonstrated conflicting evidence regarding the effect of lockdown on the prevalence of preterm birth (PTB).[7-16]
- In Australia, PTB is associated with multiple pregnancy, tobacco smoking, living in remote areas, indigenous mothers, and mothers aged ≤20 years or ≥40 years.[38]
- PTB is the leading cause of death in children under five[1], and the prevalence in Australia has remained largely static, despite perinatal care advances.[2]

## What this study adds

- During the COVID-19 lockdown and post-lockdown periods, the prevalence of moderate to late PTB decreased at the Sunshine Coast University Hospital in Queensland, Australia.
- Decreased transmission of infections[35], increased influenza vaccination rates[34], and improved air quality[24] may have been favourable in reducing PTB during the COVID-19 lockdown.
- This study highlights the need for further research into factors which may have contributed to the reduction in PTB observed during the pandemic.

## TABLE AND FIGURE CAPTION LIST

**Supplementary table 1**. Univariate analysis: logistic regression using term (0=preterm, 1=term) as the dependent (outcome) variable at the Sunshine Coast University Hospital from 2018–2020.

**Supplementary table 2**. Multivariate analysis: logistic regression using term (0=preterm, 1=term) as the dependent (outcome) variable at the Sunshine Coast University Hospital from 2018–2020.

## Notes

### Competing Interest Statement

The authors have declared no competing interest.

